# Soluble TREM2 and Alzheimer-related biomarker trajectories in the blood of diabetic patients based on their cognitive status

**DOI:** 10.1101/2022.07.07.22277348

**Authors:** Noriko Satoh-Asahara, Hajime Yamakage, Masashi Tanaka, Teruaki Kawasaki, Sayo Matsuura, Harutsugu Tatebe, Ichiro Akiguchi, Takahiko Tokuda

## Abstract

**Aim:** Type 2 diabetes mellitus (DM) increases the risk of dementia. We aimed to elucidate the dynamics of blood biomarkers according to the severity of cognitive impairment in patients with DM and to identify useful biomarkers for diabetes-related dementia.

**Methods:** This was a cross-sectional, nested case-control study of 121 Japanese diabetic and nondiabetic patients with different levels of cognitive functioning. We evaluated participants’ cognitive functions, blood biomarkers related to Alzheimer’s disease, and soluble triggering receptors expressed on myeloid cells 2 (sTREM2). We then compared these biomarkers between the DM and non-DM groups and across the different cognitive strata.

**Results:** Significantly lower levels of serum sTREM2 were observed in the DM than in the non-DM patients. This was true across all the cognitive strata of the two groups, including those with normal cognition. We also found that plasma levels of phosphorylated tau 181 (p-tau181) increased with increasing levels of cognitive decline in both the DM and non-DM groups. However, this was accompanied by a decrease in plasma amyloid-β (Aβ)42/Aβ40 ratios in non-DM patients only.

**Conclusion:** This study revealed novel characteristic trajectories of dementia-related blood biomarkers in diabetes-related dementia, suggesting the pathological involvement of molecular cascades initiated by impaired microglial activation. This results in decreased serum sTREM2, followed by tauopathy without substantial amyloid plaques, reflected by plasma p-tau elevation with no decrease in the Aβ42/Aβ40 ratio. Our results warrant further research into this molecular cascade to elucidate pathogenetic mechanisms of diabetes-related dementia and establish useful biomarkers.

## Introduction

With the unprecedented increase in the aging population, there has been a corresponding increase in the number of elderly patients with type 2 diabetes mellitus (DM) and the prevalence of dementia. Epidemiologically, DM is among the major risk factors for dementia, with more than double the risk of Alzheimer’s disease (AD) and vascular dementia (VaD) in those with DM [1, 2]. Cognitive impairment affects drug management, blood sugar control, and prognosis in patients with DM, and dementia is a serious health issue associated with disability and dependence. Hanyu suggested that a unique type of dementia affects the patients with DM, called diabetes-related dementia (DrD), which is associated with DM-related metabolic abnormalities not seen in dementia caused by typical AD or cerebrovascular pathologies [3]. Based on their studies using amyloid-positron and tau-positron emission tomography (PET), Hanyu et al. found that more than half of the patients with DrD showed positive tau deposition beginning in the medial temporal lobe, which was unaccompanied by amyloid deposition in the brain [3-5]. DM-related metabolic abnormalities such as hyperglycemia, insulin resistance, and inflammation may be implicated in the pathogenesis of DrD, but its underlying pathological mechanisms are unclear. Imaging and biofluid biomarkers that could assist in the diagnosis of DrD and its distinction from other forms of dementia have yet to be identified. Since both DM and dementia are highly prevalent disorders, blood-based biomarkers are preferable, as they are more efficient and accessible than cerebrospinal fluid (CSF) and PET imaging biomarkers.

Triggering receptor expressed on myeloid cells 2 (TREM2) is an innate immune receptor of the immunoglobulin superfamily. It is primarily expressed by monocytes, macrophages, and microglia in the central nervous system [6-8]. Genome-wide association studies have revealed an association between certain loss-of-function variants of the TREM2 gene and AD pathogenesis [9, 10]. TREM2 engagement is a key step in the microglial activation that occurs in response to pathological events in the brain, such as amyloid β (Aβ)-deposition, demyelination, and apoptotic cell death. Thus, it is involved in regulating the immune response to dementia pathologies [7, 8, 11]. TREM2 activation releases soluble TREM2 (sTREM2), the extracellular domain of the cell surface receptor, into the brain’s interstitial fluid. From there, it passes into the CSF and peripheral blood [12]. It has been suggested that sTREM2 in the CSF and blood could be a useful biomarker of microglial activity [11, 13, 14]. Higher CSF levels of sTREM2 may reflect the activation of microglia in response to neurodegeneration [15-17] and could be a preclinical indicator of AD-induced cognitive decline [17, 18]. A negative correlation has been found between serum levels of sTREM2 and cognitive function in AD patients [19]. In the general elderly Japanese population of the Hisayama study, higher levels of serum sTREM2 at baseline were found to be associated with an increased subsequent risk of both AD and VaD [20]. Furthermore, we recently demonstrated that higher levels of serum sTREM2 are cross-sectionally [21] and longitudinally [22] associated with an increased risk of diabetes-related cognitive impairment. In contrast, a recent longitudinal study of participants of the Dominantly Inherited Alzheimer Network (DIAN) found that higher CSF sTREM2 levels reduced the risk of AD pathologies [23]. These inconsistent research results make it unclear whether TREM2 plays a protective or detrimental role in the development of DrD, and whether sTREM2 could be a useful blood biomarker for early indication of the development of DrD.

Emerging evidence suggests that blood levels of Aβ, phosphorylated tau (p-tau), and neurofilament light chain (NfL), when measured using recently developed supersensitive digital enzyme-linked immuno-sorbent assays (ELISAs), could be promising biomarkers of AD pathologies [24-28]. Therefore, the present study aimed to elucidate the dynamics of dementia-related blood biomarkers in DrD based on the severity of cognitive impairment and to identify blood biomarkers that could be used to diagnose and predict the development of DrD. We examined blood levels of sTREM2 and AD biomarkers (Aβ42/Aβ40 ratio, p-tau181, total tau [t-tau], and NfL) among diabetic and non-diabetic patients with normal cognition, mild cognitive impairment (MCI), or dementia. Our findings suggested that DrD may be characterized with a cascade of events starting with DM-induced impairment of microglial activation, followed by insufficient amyloid plaque formation, which could exacerbate tau pathology resulting in neurodegeneration in the brain.

## Methods

### Study design and patients

This was a cross-sectional, nested case-control study within a cohort of patients registered with practices in an ongoing prospective, longitudinal, observational cohort study at the Kyoto Dementia Comprehensive Center Clinic (Uji, Kyoto). The base cohort study has a research database of the clinical and demographic details of more than 2,000 individuals prospectively recorded from routine clinic practice history. The data include demographic information, medical diagnoses, prescriptions, laboratory results, and clinical values. The study was approved by the Kyoto Dementia Center Research Ethics Committee and the requirement for written informed consent was waived due to the anonymization of patient data (UMIN ID: UMIN000048032). The base cohort included patients aged ≥20 years registered at the Kyoto Dementia Comprehensive Center Clinic during the study period from May 2019 to March 2020. There were no exclusion criteria.

Case patients were those with DM at the time of enrollment, with DM defined as HbA1c levels ≥6.5% or the ongoing use of diabetes medication. This definition of DM is based on the criteria of the Japan Society for the Study of Diabetes. Each case was matched to a control (nondiabetic) patient of equivalent age (within 1 year), sex, and cognitive functioning using incidence density sampling. The matching ratio was 1:2 (DM: non-DM) for normal and MCI, and 1:1 for dementia, based on the power analysis results.

### Measurement of metabolic parameters

We measured the anthropometric and metabolic parameters of all patients at baseline using standard procedures [21]. These parameters included body weight, systolic and diastolic blood pressure (SBP and DBP), heart rate, fasting plasma glucose, hemoglobin A1c (HbA1c), serum immunoreactive insulin (IRI), homeostasis model assessment of insulin resistance (HOMA-IR), triglycerides (TG), high-density lipoprotein cholesterol (HDL-C), and low-density lipoprotein cholesterol (LDL-C). SBP and DBP were measured twice with an automatic electronic sphygmomanometer (BP-103i II; Nippon Colin, Komaki, Japan). Grip strength in the dominant hand was measured with the Smedley grip force system.

### Measurement of plasma AD biomarkers

Ethylenediaminetetraacetic acid (EDTA) plasma samples were obtained via venipuncture. Plasma levels of the AD biomarkers—Aβ42, Aβ40, total tau (t-tau), p-tau181, and NfL—were quantified using a supersensitive automated digital ELISA platform (Simoa™ HD-1 analyzer, Quanterix, Lexington, KY, USA) with validated assay kits. Procedures were conducted according to the manufacturer’s instructions. Based on previous studies, we employed the plasma Aβ42/Aβ40 ratio as an indicator of brain amyloid burden [25, 26] instead of the Aβ42 levels that are widely used as a CSF biomarker for brain amyloid.

### Measurement of serum sTREM2

Serum sTREM2 levels were quantified at Health Science West Japan (Kyoto, Japan) using a RayBiotech Human TREM-2 ELISA Kit (RayBiotech, Norcross, GA, USA), according to the manufacturer’s instructions and as described in a previous study [21]. The patients were divided into four categories according to their serum sTREM2 levels.

### Assessment of cognitive function

#### MMSE

We assessed cognitive function using the Japanese version of the Mini-Mental State Examination (MMSE) (Nihon Bunka Kagakusha Co. Ltd., Tokyo [provided by Psychological Assessment Resources, Inc., FL, USA]). Possible scores on the MMSE range from 0 to 30, with higher scores indicating better cognitive function. We classified participants with MMSE scores of 27–30 as normal, those with scores of 24–26 as MCI, and those with scores ≤23 as having dementia [29]. These classifications were based on the guidelines of the Japanese Society of Neurology (https://www.neurology-jp.org/guidelinem/nintisyo_2017.html), which was developed in cooperation with other Japanese societies related to dementia.

#### DASC-21

The Dementia Assessment Sheet for the Community-based Integrated Care System (DASC-21) comprises 21 questions, each scored on a 4-point scale, allowing a total score range of 21–84 [30]. A higher score indicates greater cognitive impairment. Scores of >31 indicate a risk for dementia, and a score of 3 or 4 for an individual item indicates impairment in that aspect of cognition. DASC-21 scores are significantly correlated with those of the Clinical Dementia Rating scale [30].

### Statistical analysis

The sample size required to detect valid differences in effect size (d = 0.8) between groups was calculated for a significance level of 0.05 and a power of 80%. The DM group was selected from the original cohort using the inclusion criteria of this study, and the non-DM group was selected with an allocation ratio that ensured power.

Data were described as means and standard deviations (SD) or as medians and interquartile ranges (IQR). A generalized linear model with a logarithmic link function was used for comparing the sTREM2 levels and AD biomarkers between groups. Fixed factors included DM status (DM or non-DM), cognitive functioning (normal, MCI, or dementia), and the interactions between these groups (DM × dementia). Statistics were calculated as geometric means and their 95% confidence intervals (CIs). Spearman’s rank correlation coefficient (ρ) tests were employed to investigate the correlations of sTREM2 and AD biomarkers with anthropometric parameters, metabolic parameters, and the levels of cognitive function. A two-sided *p*-value <0.05 was considered statistically significant. All statistical analyses were performed using SPSS for Windows version 24.0 (IBM Corp., Armonk, NY, USA) software.

## Results

### Baseline characteristics

A total of 121 participants were included in this study. The characteristics of patients in the DM groups (n = 47; 14 patients with normal cognition, 13 with MCI, and 20 with dementia) are shown in Table 1, and those of the non-DM groups (n = 74; 28 participants with normal cognition, 26 with MCI, and 20 with dementia) in Table 2. The DM and non-DM groups were matched for age and gender (mean age range: 78.9–80.6 years; male/female ratio of each cognitive group: normal, 1:2, MCI: 1:2, dementia: 1:1). The MMSE scores were also confirmed to be balanced between the DM and non-DM groups (normal: DM, 28.4 ± 1.0 and non-DM, 28.3 ± 1.1; MCI: 25.1 ± 0.9 and 25.0 ± 0.8; dementia: 17.3 ± 3.6 and 18.1 ± 3.8).

**Table 1.**
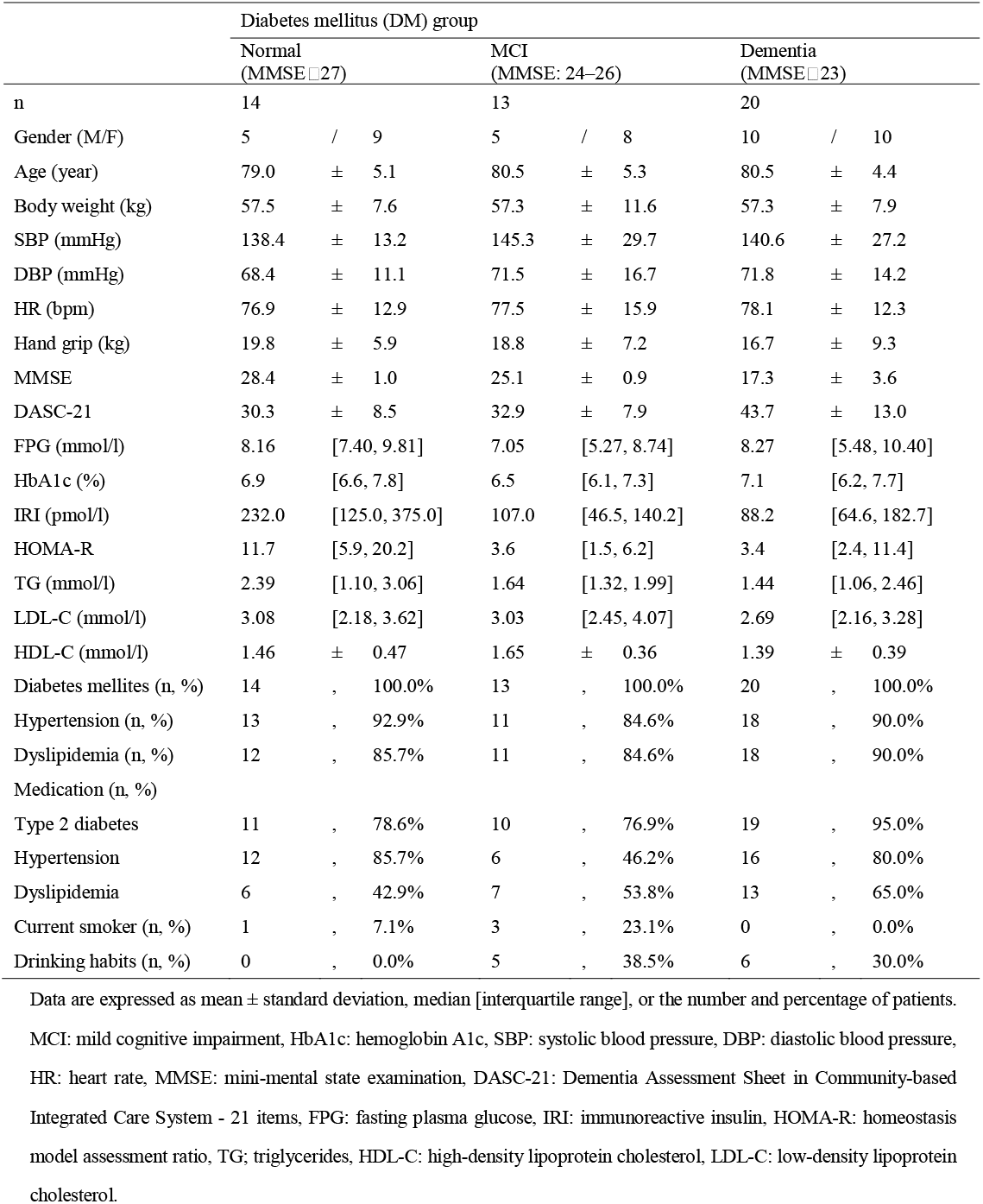
Baseline characteristics of the patients with diabetes mellites (DM group)

**Table 2.** Baseline characteristics of the patients without diabetes mellites (non-DM group)

|  | Non-diabetes mellitus (non-DM) group |  |  |  |  |  |  |  |  |
| --- | --- | --- | --- | --- | --- | --- | --- | --- | --- |
| | Normal<br>(MMSE ≥ 27) | | | MCI<br>(MMSE: 24–26) | | | Dementia<br>(MMSE ≤ 23) | | |
| n | 28 |  |  | 26 |  |  | 20 |  |  |
| Gender (M/F) | 10 | / | 18 | 10 | / | 16 | 10 | / | 10 |
| Age (year) | 78.9 | ± | 5.3 | 79.4 | ± | 5.1 | 80.6 | ± | 4.6 |
| Body weight (kg) | 54.7 | ± | 9.0 | 52.6 | ± | 9.4 | 52.4 | ± | 12.0 |
| SBP (mmHg) | 144.5 | ± | 20.1 | 137.1 | ± | 28.6 | 142.1 | ± | 27.0 |
| DBP (mmHg) | 75.7 | ± | 11.5 | 71.8 | ± | 15.6 | 78.3 | ± | 14.0 |
| HR (bpm) | 75.9 | ± | 12.2 | 79.4 | ± | 15.5 | 83.6 | ± | 17.7 |
| Hand grip (kg) | 20.0 | ± | 6.6 | 18.6 | ± | 7.1 | 15.7 | ± | 5.7 |
| MMSE | 28.3 | ± | 1.1 | 25.0 | ± | 0.8 | 18.1 | ± | 3.8 |
| DASC-21 | 25.5 | ± | 4.8 | 29.0 | ± | 6.9 | 43.6 | ± | 12.3 |
| FPG (mmol/l) | 4.86 | [4.35, 5.76] |  | 4.69 | [4.19, 5.49] |  | 4.66 | [4.22, 5.76] |  |
| HbA1c (%) | 5.7 | [5.4, 5.9] |  | 5.5 | [5.3, 5.9] |  | 5.4 | [5.3, 5.8] |  |
| IRI (pmol/l) | 64.6 | [42.4, 172.9] |  | 59.0 | [31.3, 100.0] |  | 56.3 | [21.5, 101.4] |  |
| HOMA-R | 1.5 | [1.3, 6.2] |  | 2.0 | [1.0, 3.5] |  | 1.6 | [0.5, 3.2] |  |
| TG (mmol/l) | 1.25 | [0.81, 2.10] |  | 1.05 | [0.76, 1.67] |  | 0.96 | [0.62, 1.42] |  |
| LDL-C (mmol/l) | 2.77 | [2.34, 3.27] |  | 3.26 | [2.74, 3.83] |  | 3.10 | [2.18, 3.88] |  |
| HDL-C (mmol/l) | 1.77 | ± | 0.48 | 1.79 | ± | 0.45 | 1.77 | ± | 0.35 |
| Diabetes mellitus (n, %) | 0 | , | 0.0% | 0 | , | 0.0% | 0 | , | 0.0% |
| Hypertension (n, %) | 24 | , | 85.7% | 21 | , | 80.8% | 17 | , | 85.0% |
| Dyslipidemia (n, %) | 17 | , | 60.7% | 15 | , | 57.7% | 8 | , | 40.0% |
| Medication (n, %) |  |  |  |  |  |  |  |  |  |
| Type 2 diabetes | 0 | , | 0.0% | 0 | , | 0.0% | 0 | , | 0.0% |
| Hypertension | 16 | , | 57.1% | 16 | , | 61.5% | 8 | , | 40.0% |
| Dyslipidemia | 12 | , | 42.9% | 7 | , | 26.9% | 4 | , | 20.0% |
| Current smoker (n, %) | 3 | , | 10.7% | 2 | , | 7.7% | 4 | , | 20.0% |
| Drinking habits (n, %) | 9 | , | 32.1% | 8 | , | 30.8% | 6 | , | 30.0% |

Data are expressed as mean ± standard deviation, median [interquartile range], or the number and percentage of patients. non-DM, non-type 2 diabetes mellitus; Other abbreviations used in this table are the same as in Table 1.

### Comparison of the blood biomarker levels of DM and non-DM patients and between patients with differing levels of cognitive function

Figure 1 compares the levels of the blood biomarkers evaluated between the six groups categorized by DM status and level of cognitive function.

**Fig. 1.**
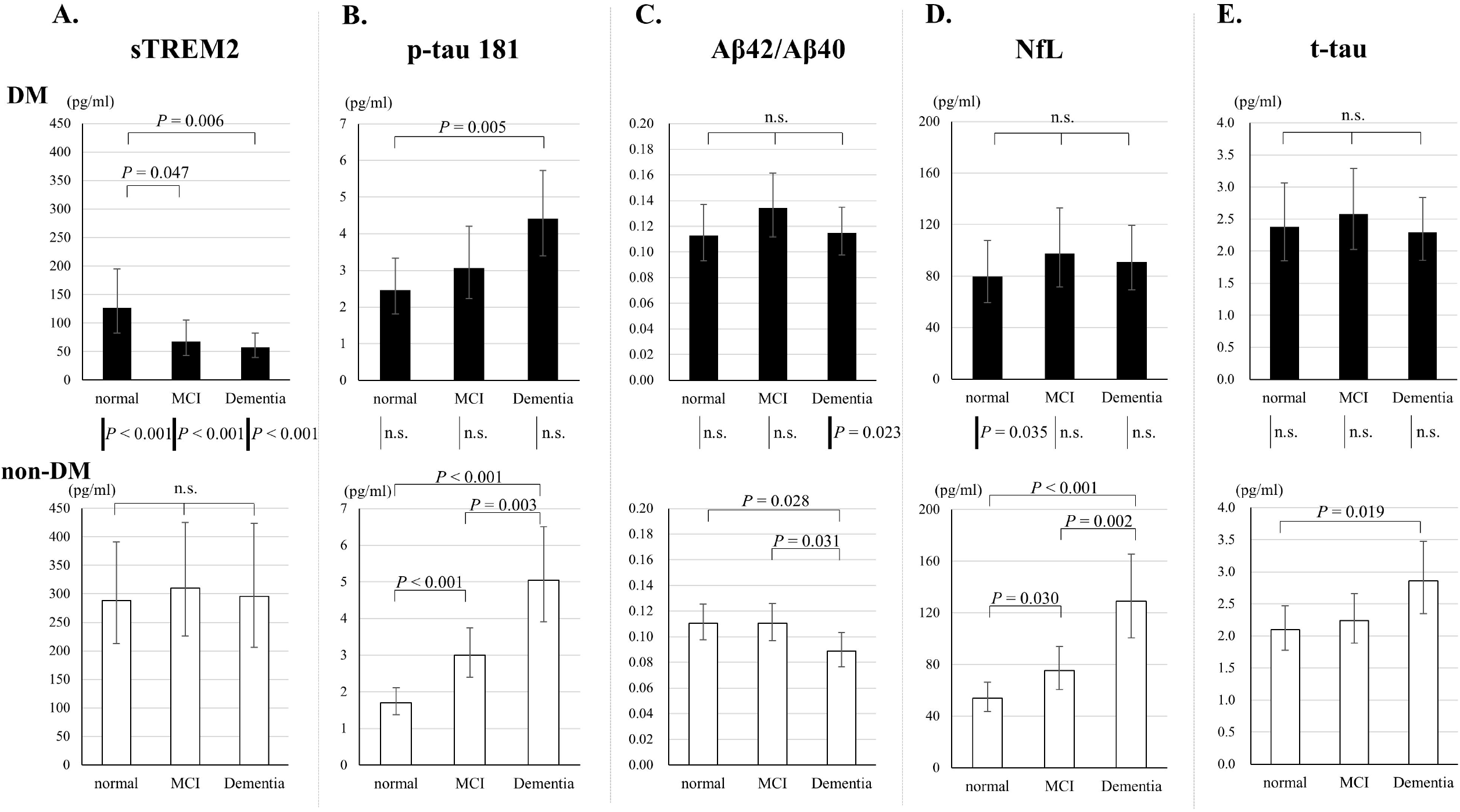
Comparison of blood biomarker levels among the six groups classified according to the type 2 diabetes myelitis (DM) status and cognitive status (normal, mild cognitive impairment [MCI], and dementia). A) Soluble triggering receptors expressed on myeloid cells 2 (sTREM2); B) Phosphorylated tau (p-tau) 181; C) Plasma amyloid-β (Aβ42/Aβ40); D) Neurofilament light chain (NfL); E) Total tau (t-tau). A generalized linear model with a logarithmic link function was used for group comparisons of biomarkers. Bars indicate geometric means; error bars indicate 95% confidence intervals. *P* values are shown for comparisons between cognitive status and between DM and non-DM. n.s = not significant.

The most remarkable finding in the DM groups (Figure 1, upper row) was the decrease in serum levels of sTREM2 corresponding to increasing cognitive impairment (*p* < 0.05), with significantly lower sTREM2 in the MCI and dementia groups than the normal group in pairwise comparisons (normal: geometric mean = 126.8 [95%CI: 82.4, 195.0]; MCI: 67.6 [43.2, 105.6], *p* = 0.047 [vs. normal]; dementia: 57.4 [40.1, 82.3] pg/mL, *p* = 0.006 [vs. normal]) (Figure 1A). In the DM groups, we also observed a trend showing increase in the plasma levels of p-tau181 with an increase in cognitive decline (*p* < 0.05), similarly to the results of the non-DM groups, and significantly higher p-tau181 levels were observed in the dementia groups than the normal group in pairwise comparisons (normal: 2.459 [1.813, 3.335]; MCI: 3.064 [2.233, 4.203]; dementia: 4.410 [3.395, 5.728] pg/mL, *p* = 0.005 [vs. normal]) (Figure 1B). However, the increase in plasma p-tau181 in the cognitively impaired DM groups was not accompanied by any significant differences in the plasma Aβ42/Aβ40 ratios between groups with different cognitive statuses (normal: 0.113 [0.093, 0.137]; MCI: 0.134 [0.111, 0.161]; dementia: 0.115 [0.098, 0.135]) (Figure 1C), in contrast to the results of the non-DM groups. There were no significant differences in NfL or t-tau between the DM cognitive function groups (NfL: normal: 79.8 [59.2, 107.5]; MCI: 97.5 [71.6, 69.4]; dementia: 90.9 [69.4, 119.2] pg/ml) (Figure 1D); (t-tau: normal: 2.380 [1.848, 3.064]; MCI: 2.579 [2.023, 3.288]; dementia: 2.294 [1.855, 2.837] pg/ml) (Figure 1E).

In the non-DM groups (Figure 1, lower row), there were no significant differences in the serum sTREM2 levels of the non-DM groups with different cognitive status (normal: 288.7 [212.9, 391.3]; MCI: 309.9 [225.9, 424.9]; dementia: 295.7 [206.3, 423.8] pg/mL) (Figure 1A) in contrast to the results of the DM groups. Further, we found that the trajectories of AD plasma biomarkers were similar to those reported in previous studies of patients with AD continuum. The p-tau181 levels significantly increased and the Aβ42/ Aβ40 ratios significantly decreased, with worsening cognitive functions (*p* < 0.05) (Figures 1B and 1C). In pairwise comparisons, p-tau181 was significantly higher in the MCI group than the normal group, and it was higher in the dementia group than the MCI group (normal: 1.703 [1.373, 2.112]; MCI: 2.998 [2.397, 3.749], p < 0.001 [vs. normal]; dementia: 5.043 [3.908, 6.507] pg/mL, p < 0.001 [vs. normal], p = 0.003 [vs. MCI]) (Figure 1B). The Aβ42/Aβ40 ratios were significantly lower in the dementia group than in the normal and MCI groups (normal: 0.111 [0.097, 0.125]; MCI: 0.111 [0.097, 0.126]; dementia: 0.089 [0.077, 0.103], p = 0.028 [vs. normal], 0.031 [vs. MCI]) (Figure 1C). The plasma levels of NfL were significantly higher in the non-DM MCI group than in the non-DM normal group, and they were also significantly and progressively higher in the dementia group than in the MCI group (normal: 53.8 [43.6, 66.4]; MCI: 75.4 [60.6, 93.9], *p* = 0.030 [vs. normal]; dementia: 128.9 [100.4, 165.4] pg/ml, *p* <0.001 [vs. normal], *p* = 0.002 [vs. MCI]) (Figure 1D). Plasma levels of t-tau were higher in the non-DM dementia group than in the non-DM normal group (normal: 2.096 [1.777, 2.474]; MCI: 2.238 [1.885, 2.657]; dementia: 2.856 [2.348, 3.474] pg/mL, *p* = 0.019 [vs. normal]) (Figure 1E).

Next, we compared the blood biomarkers of patients with and without DM stratified by cognitive status (normal, MCI, and dementia). The most notable result was significantly lower serum levels of sTREM2 in the DM than in the non-DM groups across all cognitive strata (*p* < 0.001 for all cognitive strata) (Figure 1A). There were no significant group differences in the plasma levels of p-tau181 and t-tau between the three cognitive strata (Figures 1B and 1E). However, the dementia DM group showed significantly higher plasma Aβ42/Aβ40 ratios than the dementia non-DM group (*p* = 0.023) (Figure 1C), and the normal DM group showed significantly higher NfL levels than the normal non-DM group (*p* = 0.035) (Figure 1D).

### Correlation analyses

In the DM group (Table 3), the only significant correlation found between blood biomarkers was a positive correlation between the plasma levels of NfL and t-tau (*p* = 0.003). The plasma Aβ42/Aβ40 ratios were not significantly correlated with any other parameters. A significant negative correlation was found between plasma levels of p-tau181 and both MMSE scores and HOMA-IR values (MMSE: *p* = 0.011; HOMA-IR: *p* = 0.049). Serum levels of sTREM2 were positively correlated with age (*p* = 0.001) and negatively correlated with serum TG and HDL-C levels (TG: *p* = 0.009; HDL-C: *p* = 0.022). Plasma levels of NfL were negatively correlated with plasma levels of HbA1c (*p* = 0.016).

**Table 3.**
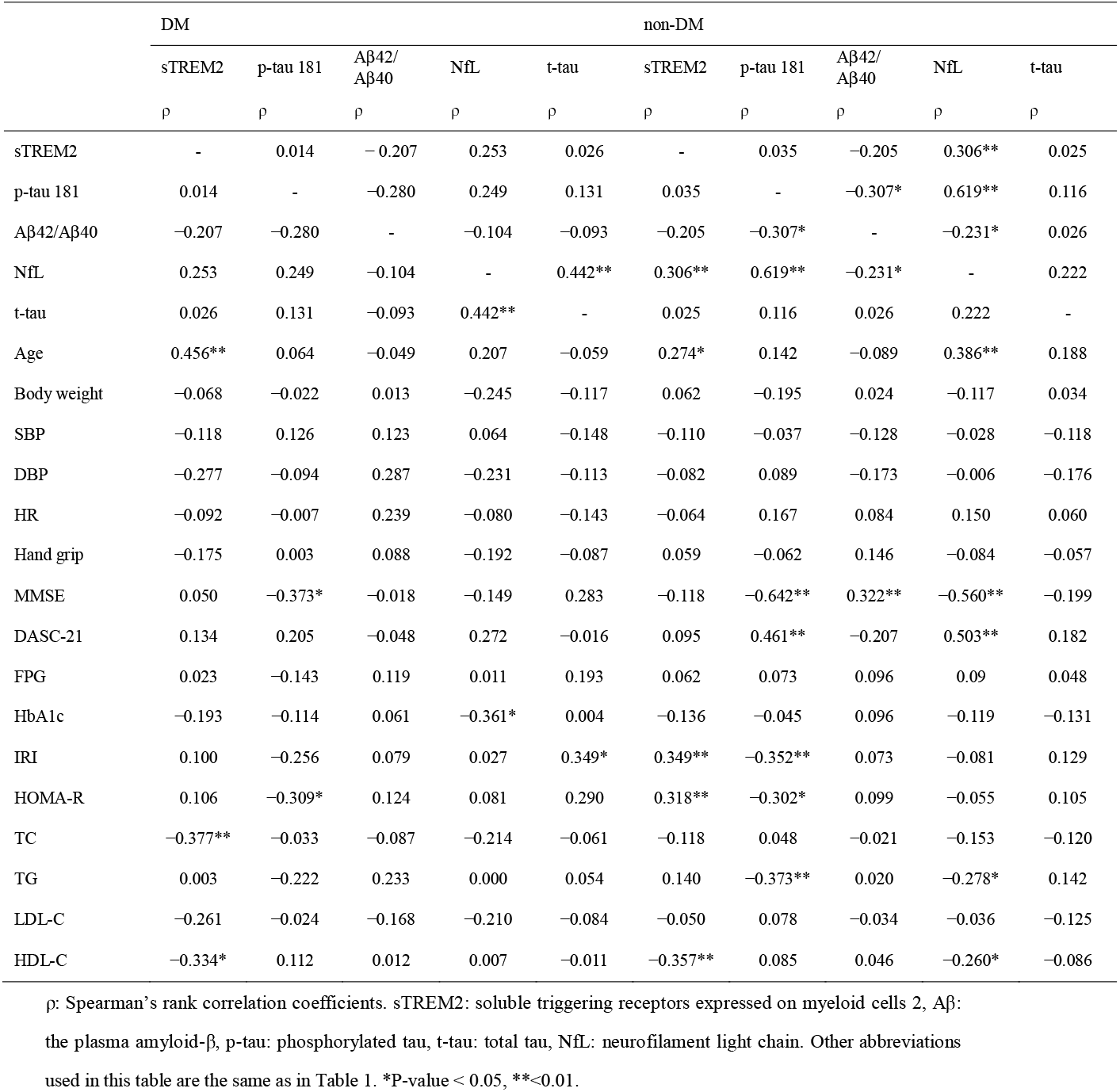
Correlation between biomarkers and glycolipid metabolism in the DM and non-DM group

|  | DM |  |  |  |  | non-DM |  |  |  |  |
| --- | --- | --- | --- | --- | --- | --- | --- | --- | --- | --- |
| | sTREM2 | p-tau 181 | Aβ42/<br>Aβ40 | NfL | t-tau | sTREM2 | p-tau 181 | Aβ42/<br>Aβ40 | NfL | t-tau |
| | $\rho$ | $\rho$ | $\rho$ | $\rho$ | $\rho$ | $\rho$ | $\rho$ | $\rho$ | $\rho$ | $\rho$ |
| sTREM2 | - | 0.014 | -0.207 | 0.253 | 0.026 | - | 0.035 | -0.205 | 0.306** | 0.025 |
| p-tau 181 | 0.014 | - | -0.280 | 0.249 | 0.131 | 0.035 | - | -0.307* | 0.619** | 0.116 |
| Aβ42/Aβ40 | -0.207 | -0.280 | - | -0.104 | -0.093 | -0.205 | -0.307* | - | -0.231* | 0.026 |
| NfL | 0.253 | 0.249 | -0.104 | - | 0.442** | 0.306** | 0.619** | -0.231* | - | 0.222 |
| t-tau | 0.026 | 0.131 | -0.093 | 0.442** | - | 0.025 | 0.116 | 0.026 | 0.222 | - |
| Age | 0.456** | 0.064 | -0.049 | 0.207 | -0.059 | 0.274* | 0.142 | -0.089 | 0.386** | 0.188 |
| Body weight | -0.068 | -0.022 | 0.013 | -0.245 | -0.117 | 0.062 | -0.195 | 0.024 | -0.117 | 0.034 |
| SBP | -0.118 | 0.126 | 0.123 | 0.064 | -0.148 | -0.110 | -0.037 | -0.128 | -0.028 | -0.118 |
| DBP | -0.277 | -0.094 | 0.287 | -0.231 | -0.113 | -0.082 | 0.089 | -0.173 | -0.006 | -0.176 |
| HR | -0.092 | -0.007 | 0.239 | -0.080 | -0.143 | -0.064 | 0.167 | 0.084 | 0.150 | 0.060 |
| Hand grip | -0.175 | 0.003 | 0.088 | -0.192 | -0.087 | 0.059 | -0.062 | 0.146 | -0.084 | -0.057 |
| MMSE | 0.050 | -0.373* | -0.018 | -0.149 | 0.283 | -0.118 | -0.642** | 0.322** | -0.560** | -0.199 |
| DASC-21 | 0.134 | 0.205 | -0.048 | 0.272 | -0.016 | 0.095 | 0.461** | -0.207 | 0.503** | 0.182 |
| FPG | 0.023 | -0.143 | 0.119 | 0.011 | 0.193 | 0.062 | 0.073 | 0.096 | 0.09 | 0.048 |
| HbA1c | -0.193 | -0.114 | 0.061 | -0.361* | 0.004 | -0.136 | -0.045 | 0.096 | -0.119 | -0.131 |
| IRI | 0.100 | -0.256 | 0.079 | 0.027 | 0.349* | 0.349** | -0.352** | 0.073 | -0.081 | 0.129 |
| HOMA-R | 0.106 | -0.309* | 0.124 | 0.081 | 0.290 | 0.318** | -0.302* | 0.099 | -0.055 | 0.105 |
| TC | -0.377** | -0.033 | -0.087 | -0.214 | -0.061 | -0.118 | 0.048 | -0.021 | -0.153 | -0.120 |
| TG | 0.003 | -0.222 | 0.233 | 0.000 | 0.054 | 0.140 | -0.373** | 0.020 | -0.278* | 0.142 |
| LDL-C | -0.261 | -0.024 | -0.168 | -0.210 | -0.084 | -0.050 | 0.078 | -0.034 | -0.036 | -0.125 |
| HDL-C | -0.334* | 0.112 | 0.012 | 0.007 | -0.011 | -0.357** | 0.085 | 0.046 | -0.260* | -0.086 |

In the non-DM group (Table 3), we found a significant negative correlation between the plasma levels of both p-tau181 and NfL and the plasma Aβ42/Aβ40 ratios (p-tau181: *p* = 0.011; NfL: *p* = 0.048). Moreover, the plasma p-tau181 and serum sTREM2 levels showed a significant positive correlation with those of NfL (p-tau181: *p* < 0.001; sTREM2: *p* = 0.008). The plasma Aβ42/Aβ40 ratios were positively correlated with MMSE scores (*p* = 0.005), and the levels of p-tau181 and NfL were negatively correlated with those (p-tau181: *p* < 0.001; NfL: *p* < 0.001). There was a significant negative correlation between plasma levels of p-tau181 and both serum IRI and HOMA-IR (IRI: *p* = 0.004; HOMA-IR: *p* = 0.013). Serum levels of sTREM2 showed a significant positive correlation with IRI and HOMA-IR (IRI: *p* = 0.004; HOMA-IR: *p* = 0.009). The levels of plasma NfL and serum sTREM2 were positively correlated with age. For lipid indices, the levels of plasma p-tau181 and NfL were negatively correlated with serum TG levels (p-tau181: *p* = 0.001; NfL: *p* = 0.016) and the levels of serum sTREM2 and plasma NfL were negatively correlated with serum HDL-C levels (sTREM2: *p* = 0.002; NfL: *p* = 0.025).

## Discussion

The present study has demonstrated, for the first time, evidence of changes in blood biomarkers associated with DrD. The following are the novel findings. 1) Most notably there were significantly lower serum levels of sTREM2 in the DM groups than in the non-DM groups across all three cognitive strata. This was significant even in the cognitively normal group and became more evident in the MCI and dementia groups. 2) In both the non-DM and DM groups, plasma p-tau181 levels increased with declining cognitive functions. However, this was accompanied by a decrease in the plasma Aβ42/Aβ40 ratios in the non-DM MCI and dementia groups, but no such association was seen in the DM groups. Based on these results, we infer that the earliest change in the biomarker trajectories associated with dementia-related brain pathologies in DM patients could be a decrease in serum sTREM2. From this, we can deduce a cascade of events in which impaired microglial activation in response to toxic brain Aβ species (reflected in the decrease in serum sTREM2) is followed by tau pathology without substantial amyloid plaques (reflected in the p-tau181 elevation with no corresponding decrease in the plasma Aβ42/Aβ40 ratio), resulting in neurodegeneration in DrD.

A further important observation was that plasma NfL levels are not a useful biomarker for neuronal damage in the brain of DM patients. In this study, plasma NfL levels were elevated even in the cognitively normal DM group, with no corresponding elevation of p-tau181. This elevation in plasma NfL was no greater in the MCI and dementia groups than in the normal group. Furthermore, plasma NfL levels were negatively correlated with plasma HbA1c levels in the DM groups. These findings suggest that increases in plasma NfL in the cognitively normal DM group are likely to result primarily from diabetic peripheral neuropathy, although plasma NfL levels in DM patients with dementia might be affected by both diabetic neuropathy and brain pathologies. Peripheral neuropathy due to DM has been reported to result in elevation of plasma NfL [31, 32]. Thus, plasma NfL is not a suitable biomarker of neuronal damage in the brains of DM patients despite its reported usefulness in this regard in a wide range of other neurological diseases, including AD [27].

In the non-DM group, the blood biomarker trajectories were roughly equivalent to those reported in the AD cohorts of previous studies [33, 34]. The serum Aβ42/Aβ40 ratios decreased in parallel with increases in plasma p-tau181 and NfL as the levels of cognitive function deteriorated. This replication of the reported trajectories of AD biomarkers supports the accuracy of our measurements of AD blood biomarkers that can evaluate AD pathologies in the brain.

In the present study, serum sTREM2 levels were found to be reduced in DM patients at all levels of cognitive functioning, including those who were cognitively normal. This suggests that microglial dysfunction induced by DM would play an important role in the pathogenesis of DrD and may begin at a preclinical stage of the disease before cognitive symptoms manifest. A recent study of mice models of AD reported that microglia phagocytose soluble Aβ species and organize compact amyloid plaques to sequester toxic Aβ species, like Aβ oligomers, by transforming into plaque-associated microglia and forming a barrier that protects brain neurons [35]. Another study created a mouse model with both AD-linked genetic backgrounds and the diabetic phenotype. These mice demonstrated impaired microglial responses to AD pathologies, with fewer microglia around Aβ-amyloid plaques and increased number of dystrophic neurites These repressive responses were found to be linked to TREM2-related processes [36]. In addition to these studies in mice, a recent longitudinal study of the DIAN cohort found that faster longitudinal increase in CSF sTREM2 correlated with slower amyloid deposition, slower cortical thinning in the precuneus, and slower decline in cognitive functions in presymptomatic AD-linked mutation carriers [23]. Taken together with these reports, our results suggest that microglial function would be impaired in the DM groups we examined since serum sTREM2 levels were lower in all the DM groups than in the respective non-DM groups. DM-induced dysfunctional microglia are unable to compact Aβ plaques, allowing the diffusion of toxic soluble Aβ species, primarily composed of Aβ42, throughout the brain. This would explain why the plasma Aβ42/Aβ40 ratios were not decreased in the DM groups. The diffusion of toxic Aβ species through the brain parenchyma due to insufficient plaque compaction would trigger tau phosphorylation in DM patients earlier than in non-DM patients as the microglia of the latter would be unimpaired. In this context, our findings suggest not only the potential protective role of microglia in the prevention of DM-related cognitive decline but also the potential utility of serum sTREM2 as a biomarker for microglial status and future risk of dementia in DM patients. In the non-DM groups, since serum sTREM2 levels were not significantly lower in the non-DM MCI and dementia groups than in the non-DM normal cognition group, we can infer that microglial function was not impaired. Thus, the unimpaired microglia could sequester toxic Aβ species, reducing plasma Aβ42/Aβ40 ratios, as is typical in AD patients.

In the DM groups, the Aβ42/Aβ40 ratios were comparable between participants with and without cognitive decline and were not significantly correlated with the plasma p-tau181 levels. These findings are consistent with those of a previous study in which amyloid-PET and tau-PET scans of patients with DrD identified not only AD-like tau deposition but also negative amyloid accumulation, which does not occur in AD [3, 4].

The pathomechanistic details of the deleterious effects of DM on microglial function remain unclear. A recent *in vitro* study found that administration of high levels of glucose resulted in microglial activation, thereby upregulating TREM2 expression and triggering inflammatory responses [37]. Since serum sTREM2 levels in this study were positively correlated with both IRI and HOMA-IR in the non-DM group, initial aggravation of glucose metabolism would trigger aberrant activation of microglia. In contrast, serum sTREM2 levels in the DM groups were lower than those in the non-DM groups, suggesting that DM impairs the microglial response to toxic Aβ species in the brain. Microglia are tissue-resident macrophages of myeloid cell origin that originate exclusively from primitive macrophages in the embryonic yolk sac [38, 39]. In peripheral tissues, impaired responses to tissue damage are a common diabetic complication. Given the central role of macrophages in the regulation of inflammation and tissue healing, it has been suggested that dysfunctional macrophages contribute to the pathogenesis of diabetic complications [40].

This study had some limitations. First, microglial function in the brain was not examined directly. Development of inexpensive *in vivo* imaging technologies to assess microglial status is required to allow such examination. Second, not all cell types capable of releasing sTREM2 into the peripheral blood have been identified. Serum sTREM2 levels appear to reflect microglial activation [20] and it has been hypothesized that adipose tissue also produces sTREM2 [22]. The comprehensive identification of sTREM2-releasing cells and further cohort studies with larger sample sizes are needed to address these issues.

In conclusion, this study provided evidence implicating DM-related microglial dysfunction in the molecular mechanisms underlying the unique pathologies observed in the brains of patients with DrD. These mechanisms appear to be cascade events initiated by an impaired microglial response to soluble toxic Aβ species and insufficient formation of amyloid plaques, with subsequent AD-like tauopathy without brain Aβ deposition. This eventually causes neuronal damage and dementia. Future studies of the pathological significance of sTREM2 will facilitate the development of predictive markers and effective treatments for cognitive decline in patients with DM.

## Data Availability

All data produced in the present work are contained in the manuscript

## Abbreviations

AD: Alzheimer’s disease
Aβ: amyloid β
CSF: cerebrospinal fluid
DASC-21: the Dementia Assessment Sheet for the Community-based Integrated Care System comprises 21 questions
DBP: diastolic blood pressure
DM: diabetes mellitus
DrD: diabetes-related dementia
FPG: fasting plasma glucose
HbA1c: hemoglobin A1c
HDL-C: high-density lipoprotein cholesterol
HOMA-IR: homeostasis model assessment ratio
hsCRP: high-sensitive C-reactive protein
IRI: immunoreactive insulin
LDL-C: low-density lipoprotein cholesterol
NfL: neurofilament light chain
MCI: mild cognitive impairment
MMSE: mini-mental state examination
PET: positron emission tomography
p-tau: phosphorylated tau
SBP: systolic blood pressure
sTREM2: soluble TREM2
TG: triglycerides
t-tau: total tau
TREM2: Triggering receptor expressed on myeloid cells 2
VD: vascular dementia.

## CRediT authorship contribution statement

**Conceptualization:** Noriko Satoh-Asahara, Ichiro Akiguchi, Takahiko Tokuda

**Methodology:** Noriko Satoh-Asahara, Hajime Yamakage, Ichiro Akiguchi

**Analysis:** Hajime Yamakage

**Investigation:** Hajime Yamakage, Sayo Matsuurae, Harutsugu Tatebe

**Resources:** Teruaki Kawasaki, Ichiro Akiguchi

**Data Curation:** Hajime Yamakage

**Writing - Original Draft:** Noriko Satoh-Asahara, Hajime Yamakage, Masashi Tanaka, Takahiko Tokuda

**Writing - Review & Editing:** Noriko Satoh-Asahara, Hajime Yamakage, Masashi Tanaka, Teruaki Kawasaki, Sayo Matsuurae, Harutsugu Tatebe, Ichiro Akiguchi, Takahiko Tokuda

**Supervision:** Noriko Satoh-Asahara

**Project administration:** Noriko Satoh-Asahara

**Funding acquisition:** Noriko Satoh-Asahara, Hajime Yamakage, Masashi Tanaka, Takahiko Tokuda

## Acknowledgments

We wish to thank Mr. Kazuya Muranaka at Kyoto Medical Center for his helpful advice and assistance. We would like to thank Satista Co., Ltd. (http://www.satista.jp/) for supporting medical statistics. We would like to thank Enago (www.enago.jp) for the English language review.

